# Journals in Aging, Geriatrics, and Gerontology: A Survey

**DOI:** 10.1101/19012278

**Authors:** Robert J. Wolff, Hannah L. Bowser

## Abstract

Aging, geriatrics, gerontology, and related areas are important areas of research as the population of older people increases in relationship to the total population. Researchers in these fields would benefit from guidance regarding sources for publishing and finding relevant scholarly journals and articles. Multiple database sites of journals were searched to provide a list of relevant publications. This list was expanded via perusal of published citation lists and searches in general search engines. A total of 243 journals were identified and examined. Of those journals, 198 journals are currently publishing and 45 journals have ceased publication. In terms of publication medium, 39% of the currently publishing journals are online-only, 4% are print-only and 59% of the journals publish both online and in print. Journals with aging in the title represent 36%, geriatrics 30% and gerontology 23%. Less than 10% have been identified as predatory journals. An expected increase in journals in the broad field of aging is indicated by the 49% of listed titles beginning in 2000 or later. This recent increase in available journals provides a need for the information listed in this paper.

## 1. Introduction

Not only is the population of elderly people rapidly increasing as a demographic, so is the number and variety of journals that publish research in the areas of aging, geriatrics and gerontology. Researchers are now finding it difficult to determine what journals they can find, which are appropriate, and how to easily access their websites.

While there are some listings of journals that can be found using search engines, none of these are very complete, nor do they provide easy access to the journals. Having a greater knowledge of, and access to, the wide variety of publishing options can make it easier for researchers and librarians to advance their professional endeavors.

The purposes of this paper include the following:

1. To document the variety of journals available in the professional areas of aging, geriatrics and gerontology.
2. To provide a list of journals that is as comprehensive as possible.
3. To help researchers and graduate students find appropriate journals in which to publish their work.
4. To open up options for other sources for publication of their research and commentaries.
5. To examine various issues in the diversity of journals in these areas.
6. To provide some warning about journals that are possibly predatory and therefore need to be avoided by serious researchers, and whose articles may be of poor quality.

## 2. Materials and Methods

The Scimago Journal website [1], the Clarivate Analytics (Web of Science) [2], JenAge Information Center [3], and PubMed (NLM) [4] were used to access lists of journals related to aging. To these we added journals found in citation listings, on publisher websites, and from internet searches. Listings of the A-Z journal titles found in the South University library system were also searched, using keywords such as “age,” “aging,” “gerontology,” etc.

The website of each journal was searched and checked for publication status and information about online and open access features, and if available, the article processing charges were determined.

Our listing of predatory journals is based on Beall’s List of Predatory Journals and Publishers, which can now be accessed in several internet sites. We used an anonymous Weebly blog [5] that was last updated 4 July 2018.

### 2.1 A Note on Predatory Publishing

Predatory journals are defined by the authors as journals that seek submissions at a high price but generally publish low quality information and often under false pretenses. Predatory journals are included in our list for three reasons. First, because they exist, it is important to list and identify them. Second, they are included so their listing can be found and avoided, by those seeking a reputable journal in which to publish. Third, we want them identified as predatory journals so that their status is clearly noted and so their published articles may be carefully analyzed as they are often of poor quality.

The problems of predatory journals and publishers have been clearly documented, though not without conflict. Warnings about predatory journals have been sounded as a threat to and an undermining of confidence in scholarly research [6-8]. Criteria are available for recognition of legitimate or predatory journals [9-11]. However, it is worth noting that the open access model of publishing does not automatically denote a predatory journal, nor are all predatory journals open access publishers and journals. Although the authors used Beall’s List as a convenient listing of predatory journals, it is important to note that this list has its critics [12,13] and that the best methods of recognizing a predatory publisher or journal are through each author’s own research [9,10].

## 3. Results

The number of currently publishing journals in the broad fields of aging, geriatrics and gerontology that is we focused on is 198, and we found 45 titles that have ceased publication. Table 1 provides basic information about the content of the journals, and the use of common terms or prefixes in journal titles. Age or Aging, with the spelling variant of Ageing, and the Portuguese Envelhecimento, were the most common descriptors used in journal titles at 70 (36%). This was followed by Geri- or Geriatric, or the Dutch Ouderengeneeskunde, with 59 (30%) uses.

**Table 1.**
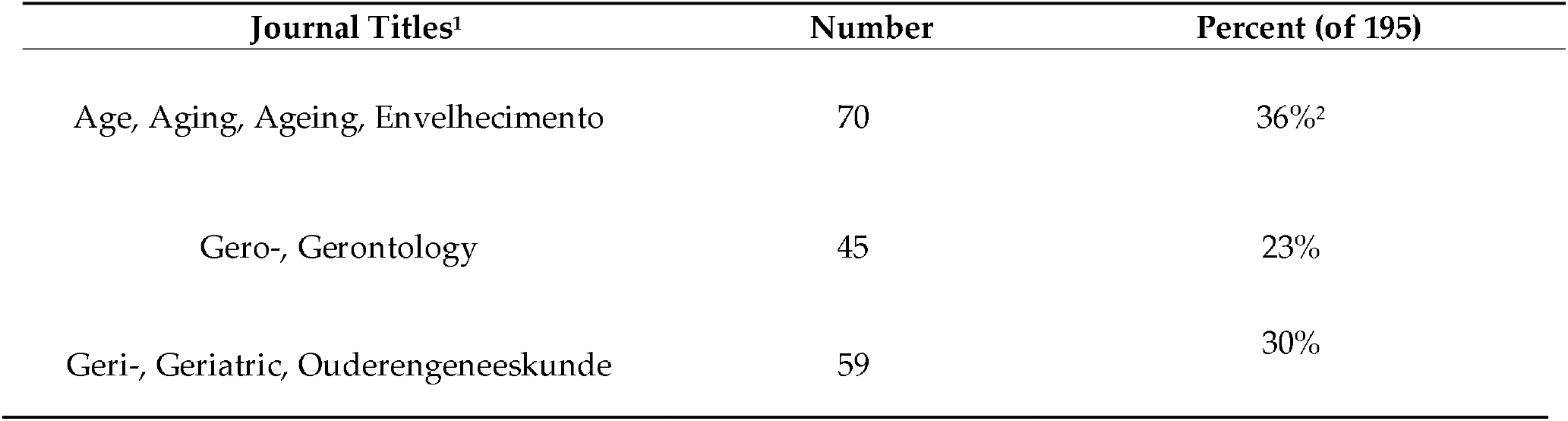
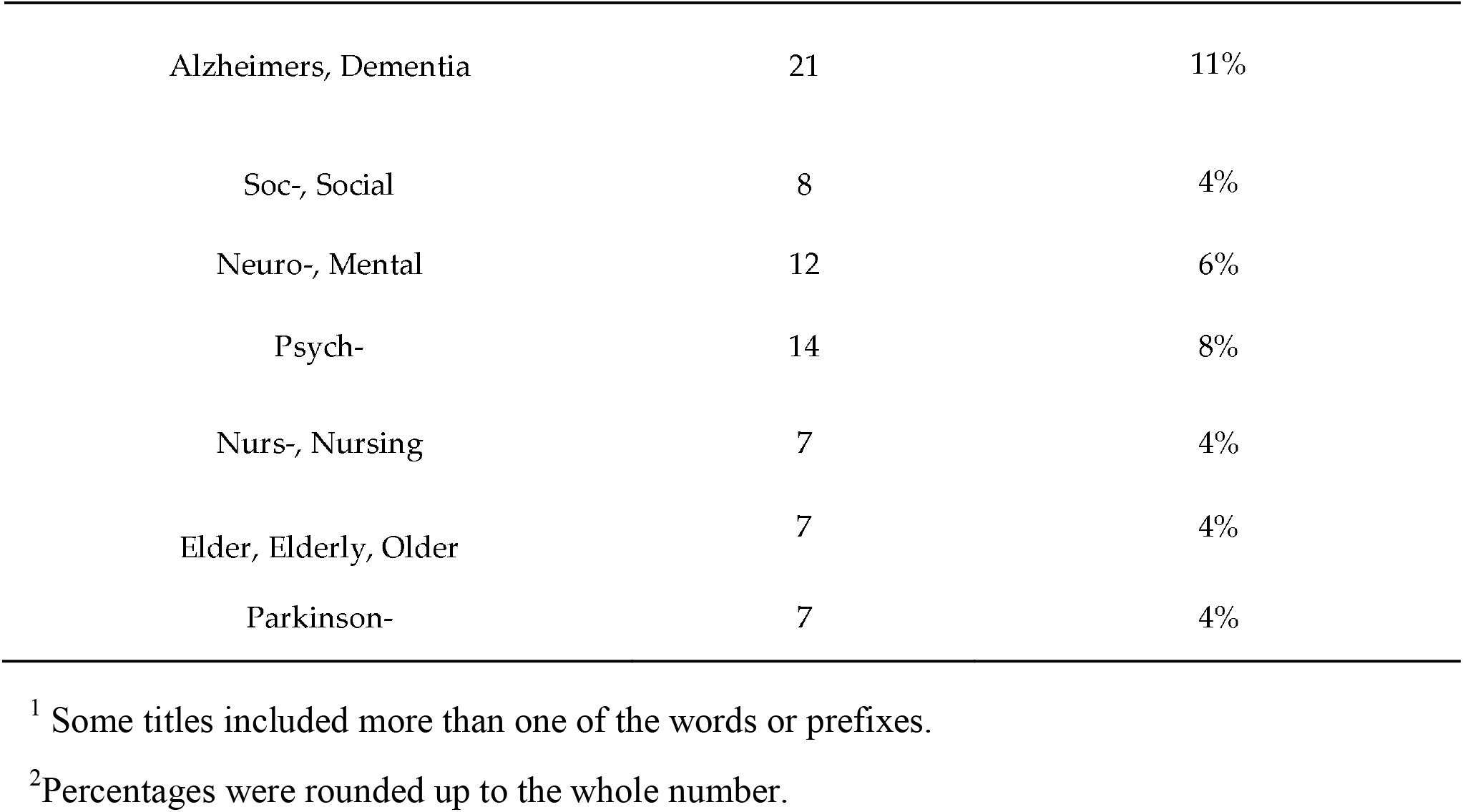
Journal areas of content (Currently publishing)

| Journal Titles<sup>1</sup> | Number | Percent (of 195) |
| --- | --- | --- |
| Age, Aging, Ageing, Envelhecimento | 70 | 36%<sup>2</sup> |
| Gero-, Gerontology | 45 | 23% |
| Geri-, Geriatric, Ouderengeneeskunde | 59 | 30% |
| Alzheimers, Dementia | 21 | 11% |
| Soc-, Social | 8 | 4% |
| Neuro-, Mental | 12 | 6% |
| Psych- | 14 | 8% |
| Nurs-, Nursing | 7 | 4% |
| Elder, Elderly, Older | 7 | 4% |
| Parkinson- | 7 | 4% |

<sup>1</sup> Some titles included more than one of the words or prefixes.
<sup>2</sup>Percentages were rounded up to the whole number.

The journals were categorized as Print (only), Online, or Both. Only 7 of the currently publishing journals were print only (4%), while 5 of the 45 journals that have ceased publication (11%) were print only, which may represent the slow demise of print journals.

Of the currently publishing journals, 76 (39%) were online only, and 115 (59%) were published both online and in print form. One journal was considered to be a print publication, with only society members having access to the online version.

Journals with 19 or less volumes published as of September 2018 numbered 98 (50%), indicating the rapid expansion of journals since 2000. Of the journals that have ceased publication, 29 (65%) began in 2000 or after, and if 1998-99 are included, the number jumps to 34 (76%), indicating the difficulty of journal start-ups to maintain publication during the move to open access publishing.

Nineteen of the currently publishing journals were identified as potentially predatory journals or publishers, just less than 10% of all available journals in the broad area of aging. We did not find any hijacked journals (where websites or branding were coopted by a predatory publisher), but the similarity of some journal names may be a form of hijacking by attempting to create confusion.

The complete list of journals that are currently being published may be found in Table 2 and the complete list of journals that have ceased publication is in Table 3.

**TABLE 2.**
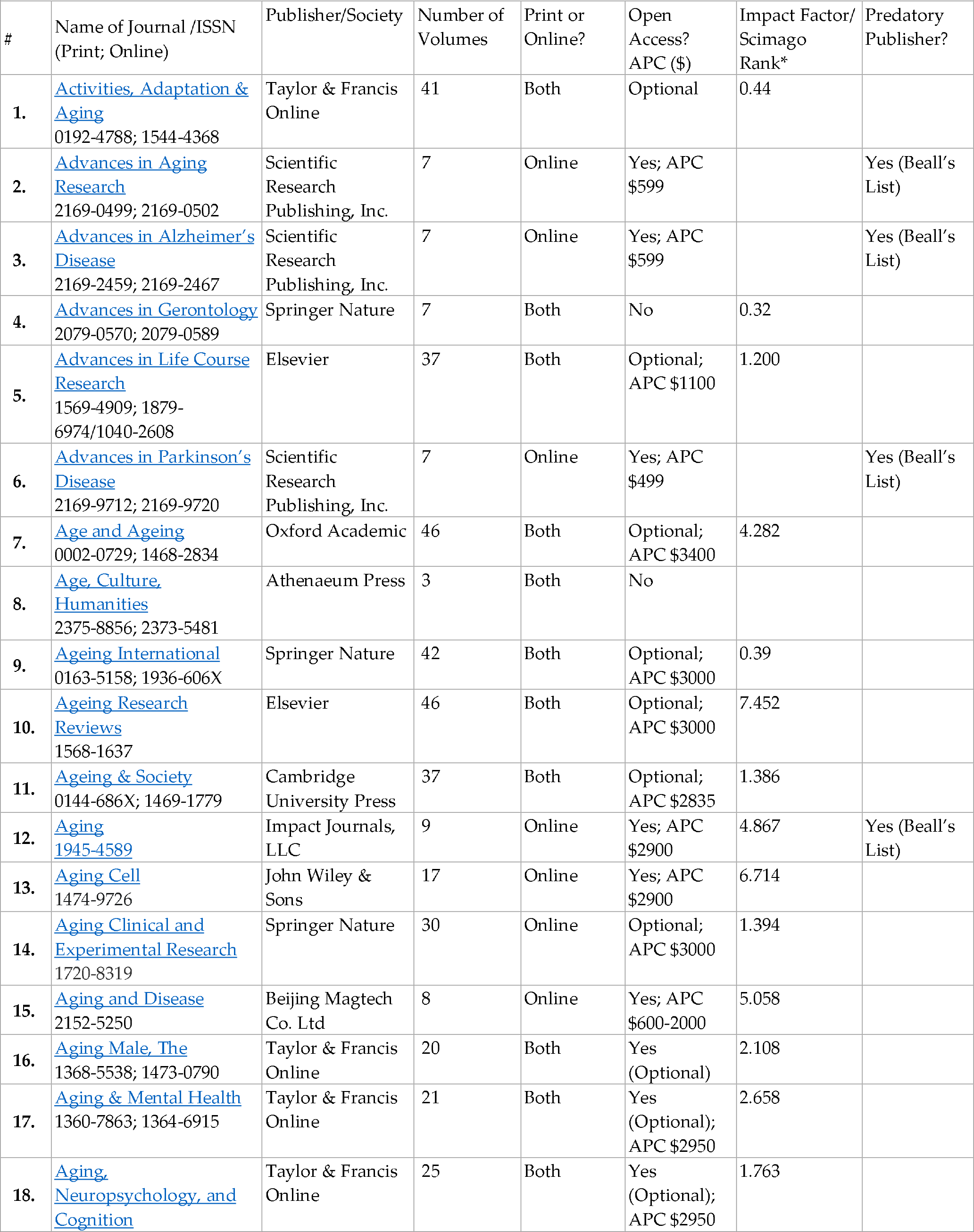
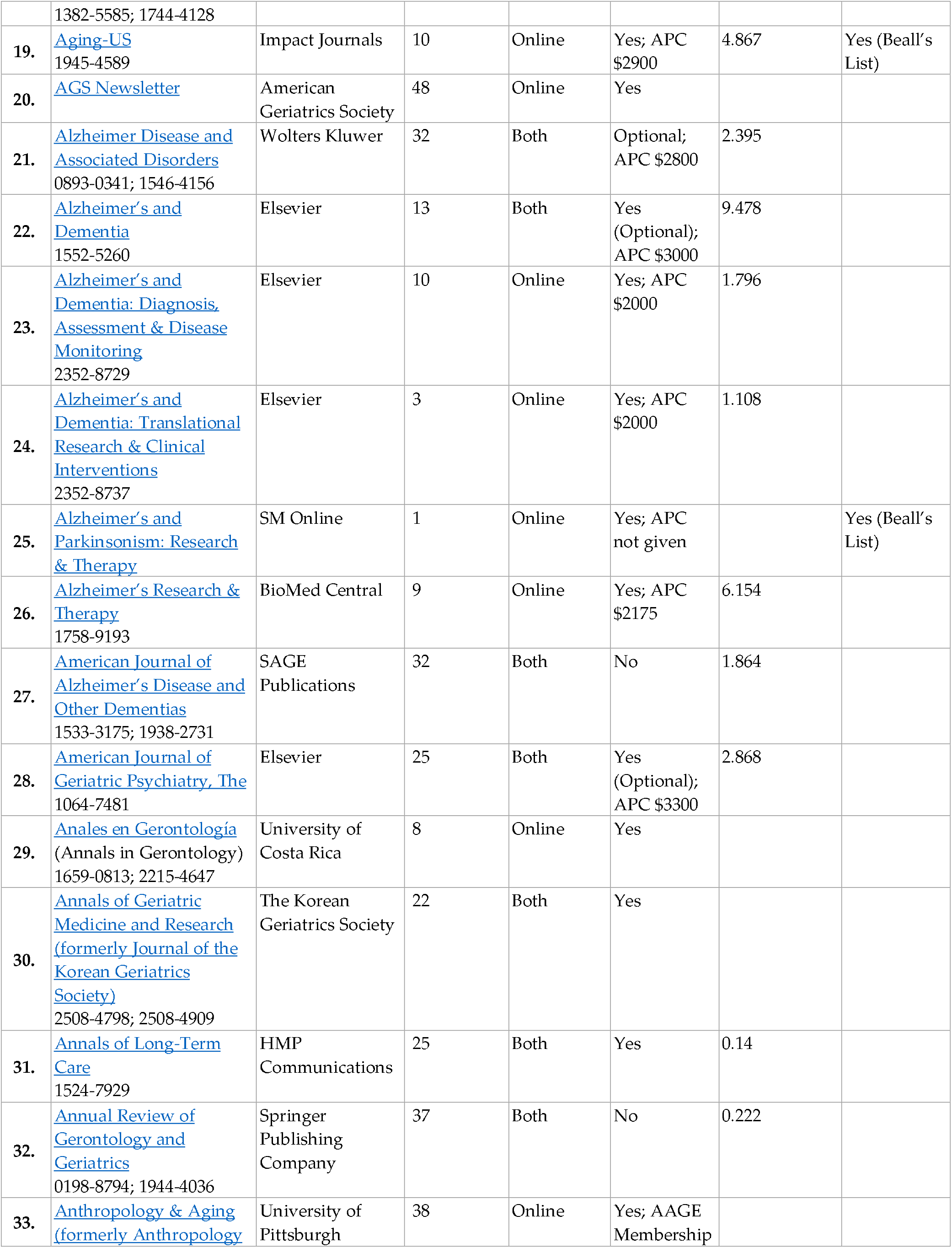
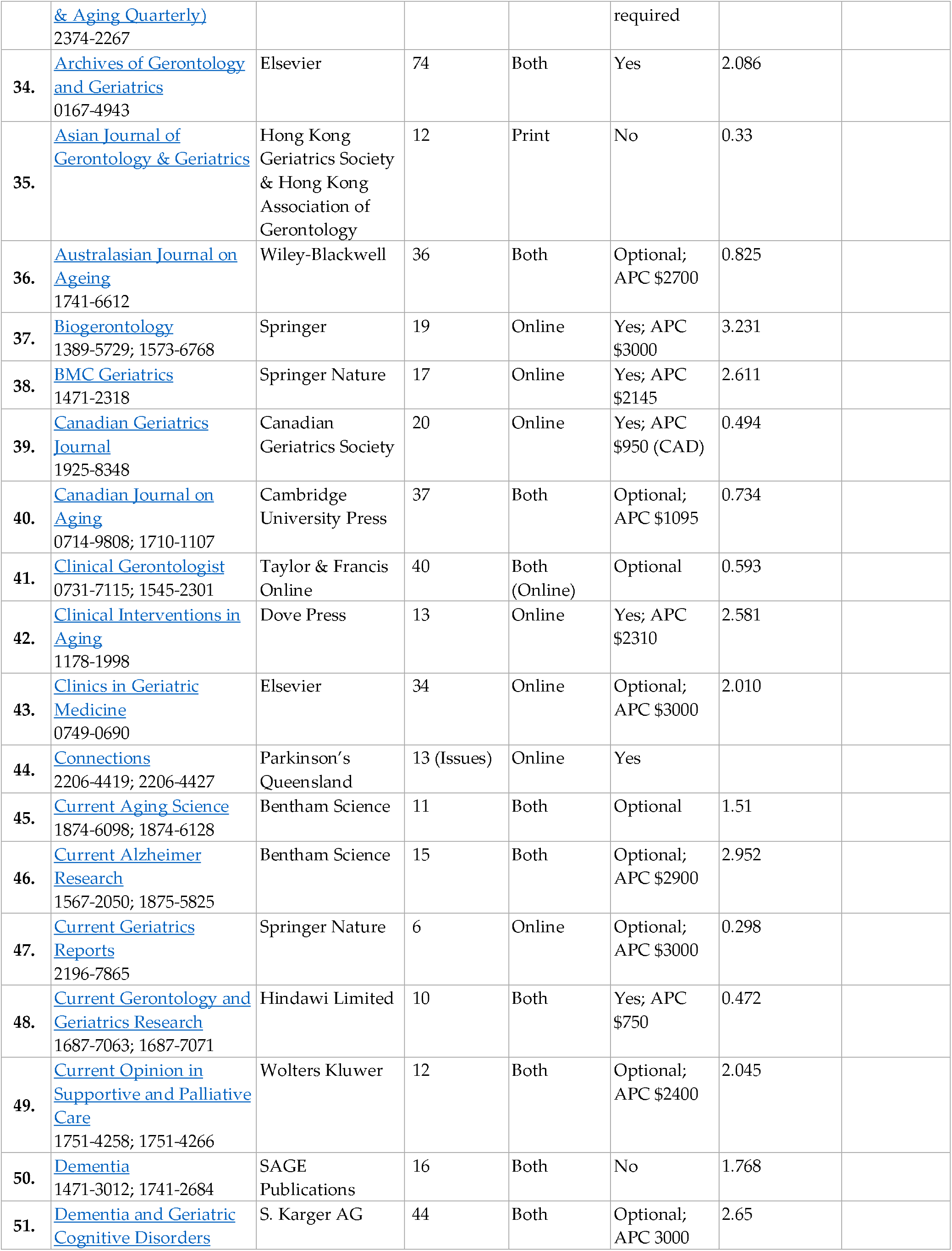
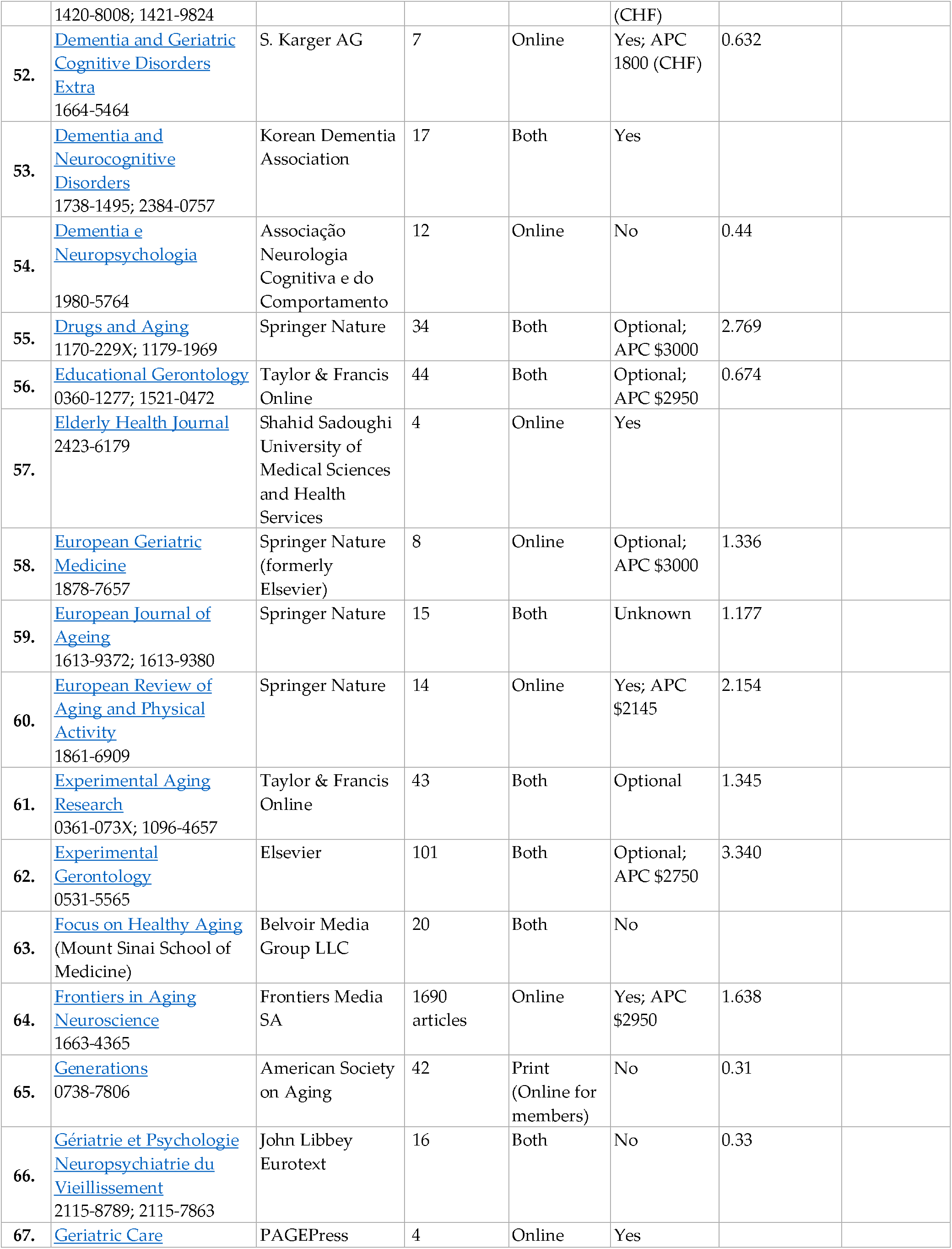
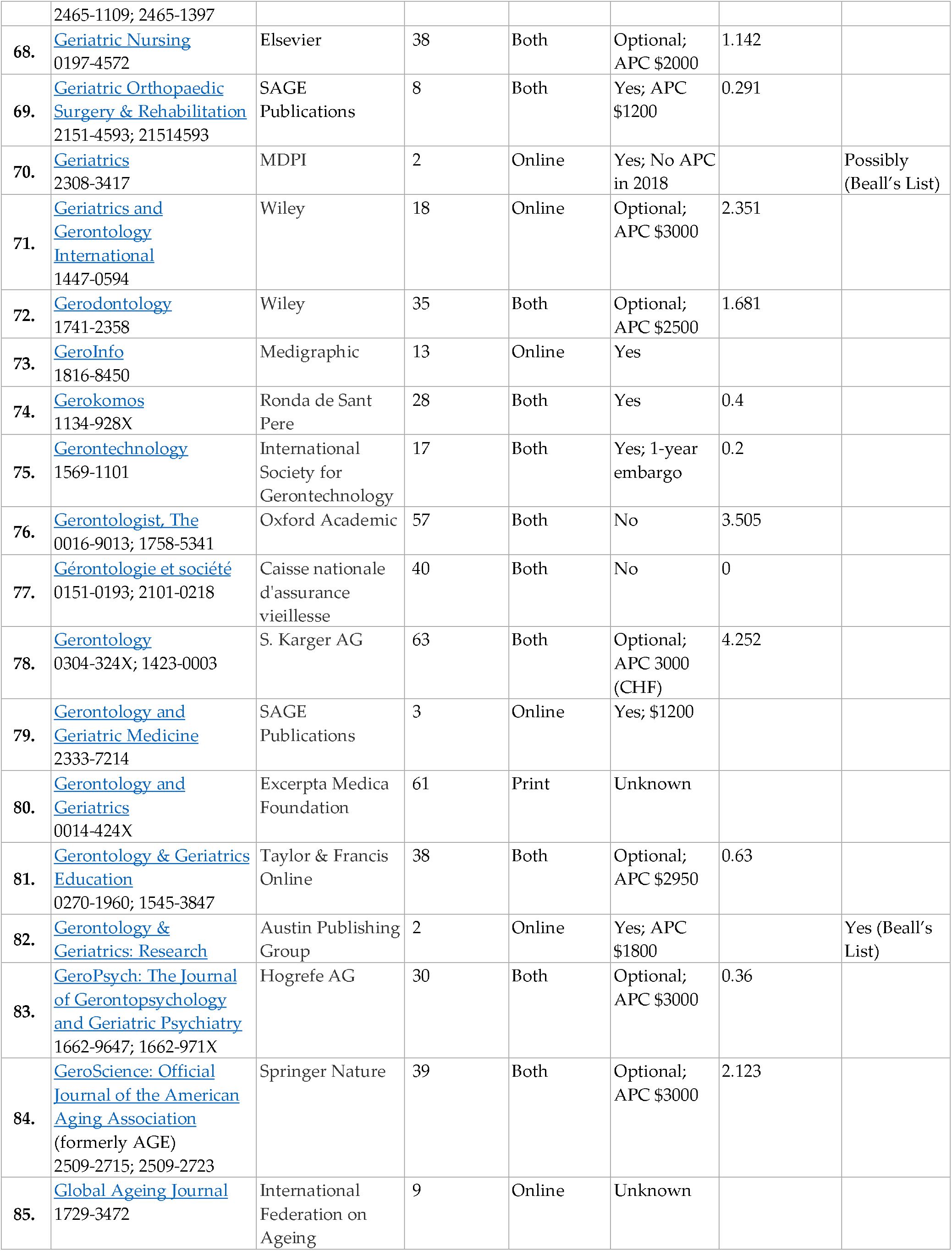
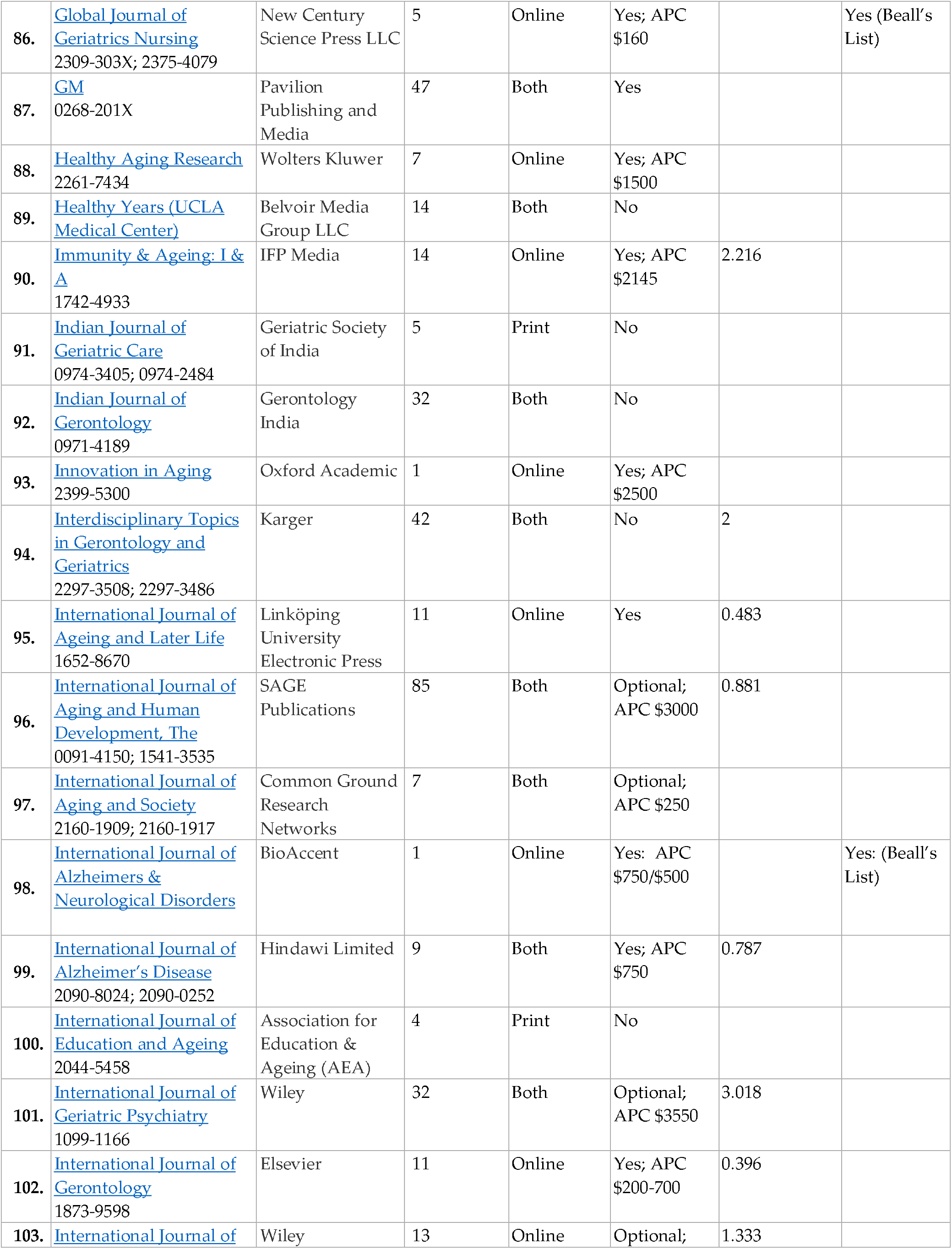
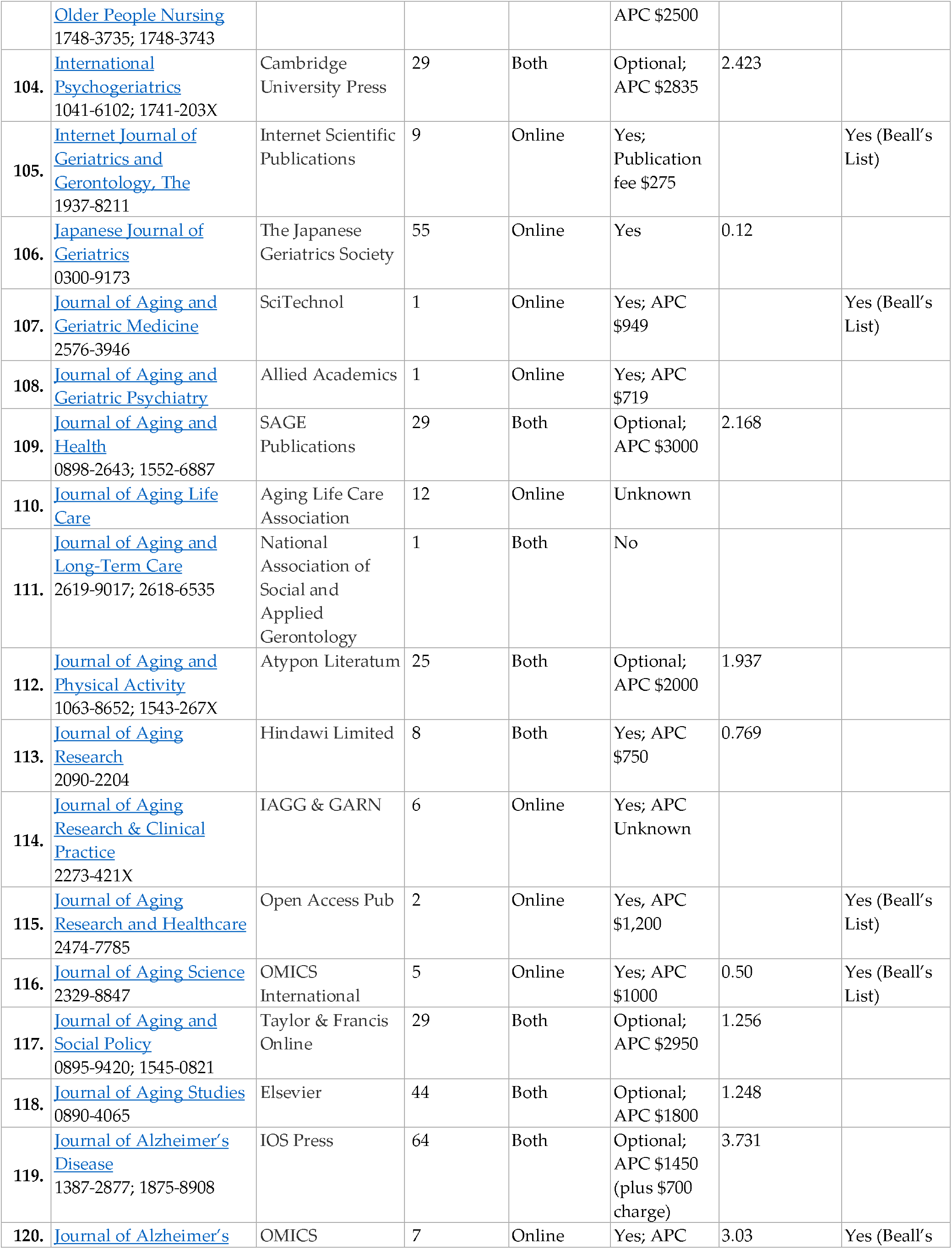
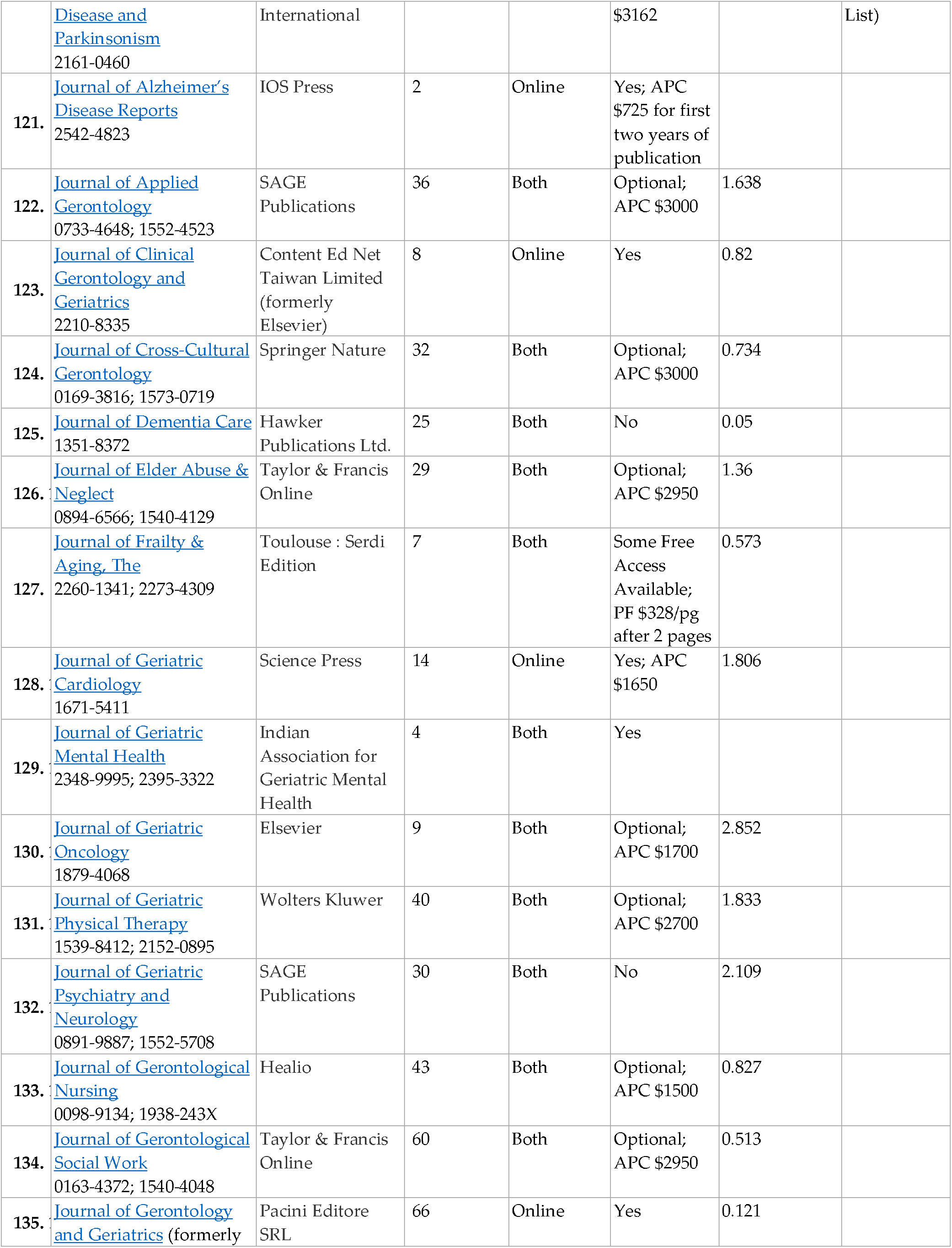
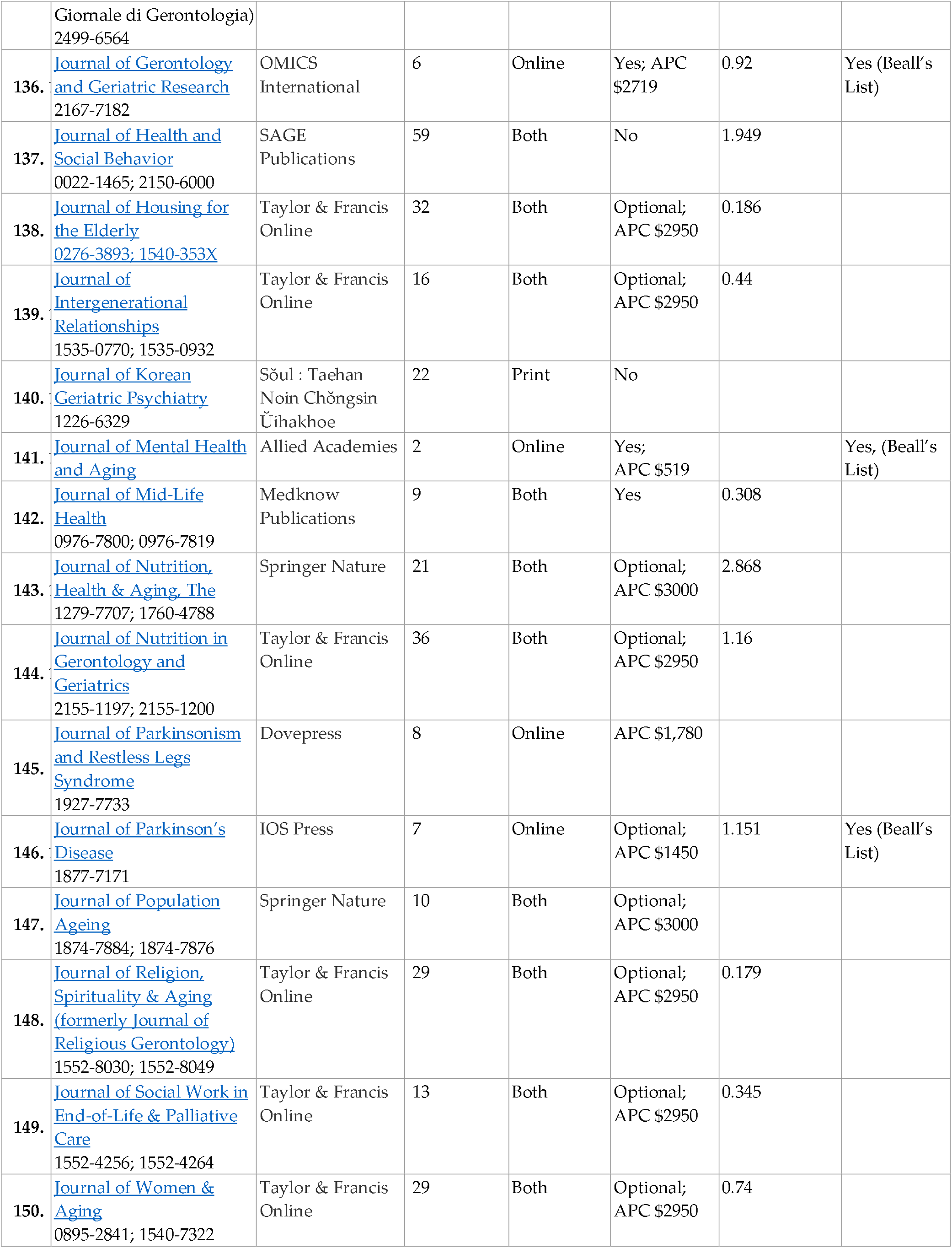
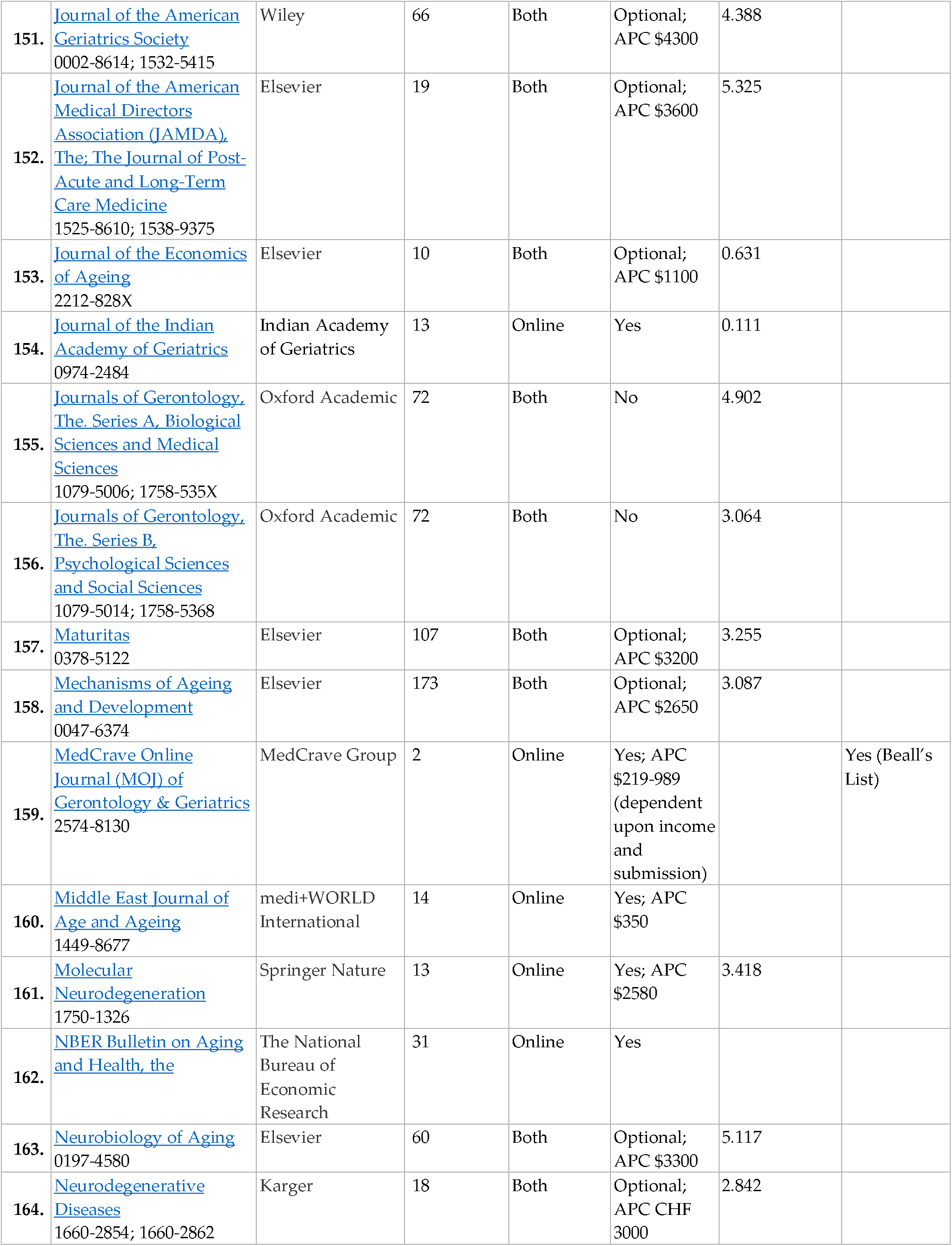
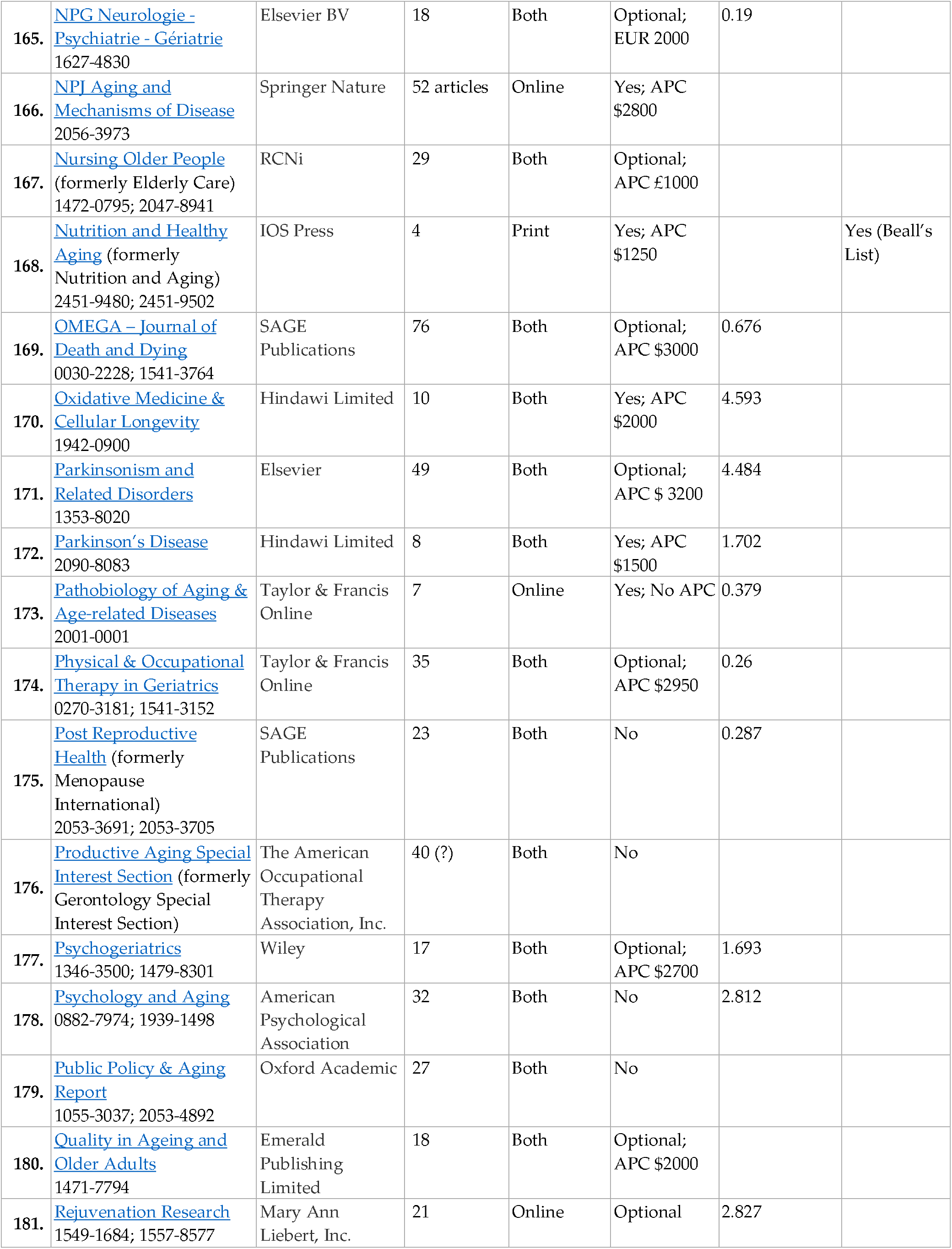
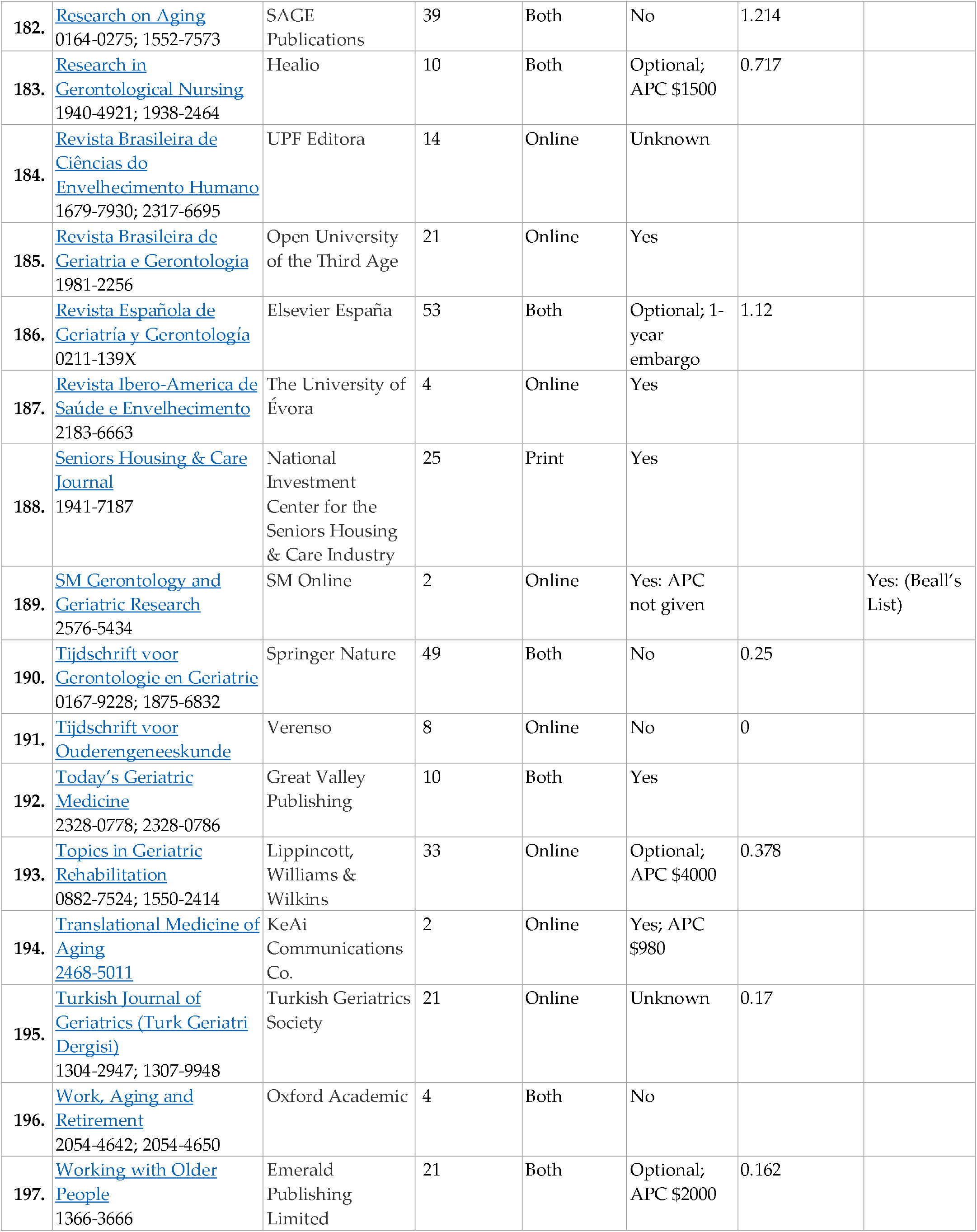
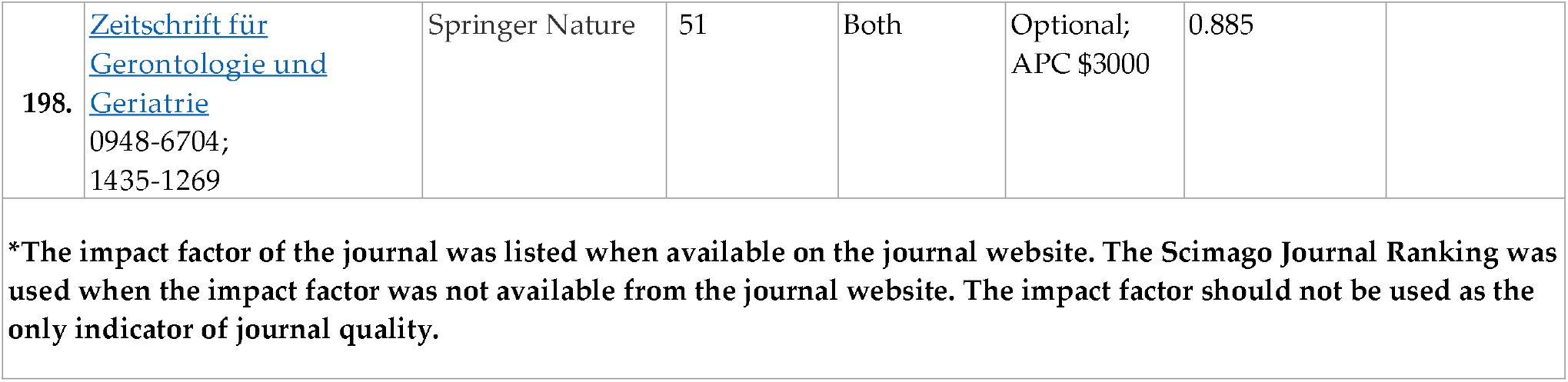
JOURNALS CURRENTLY IN PUBLICATION

**TABLE 3.**
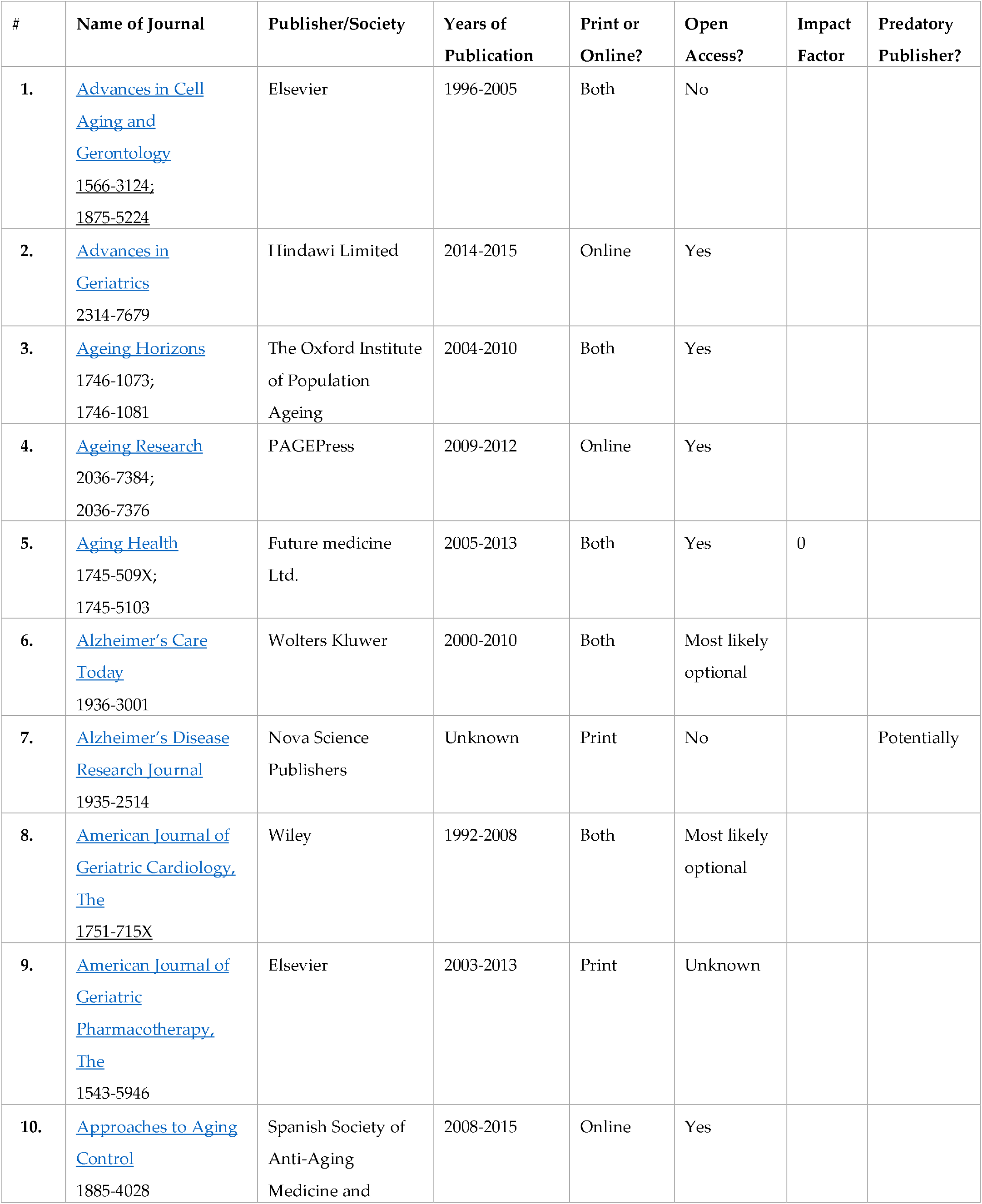
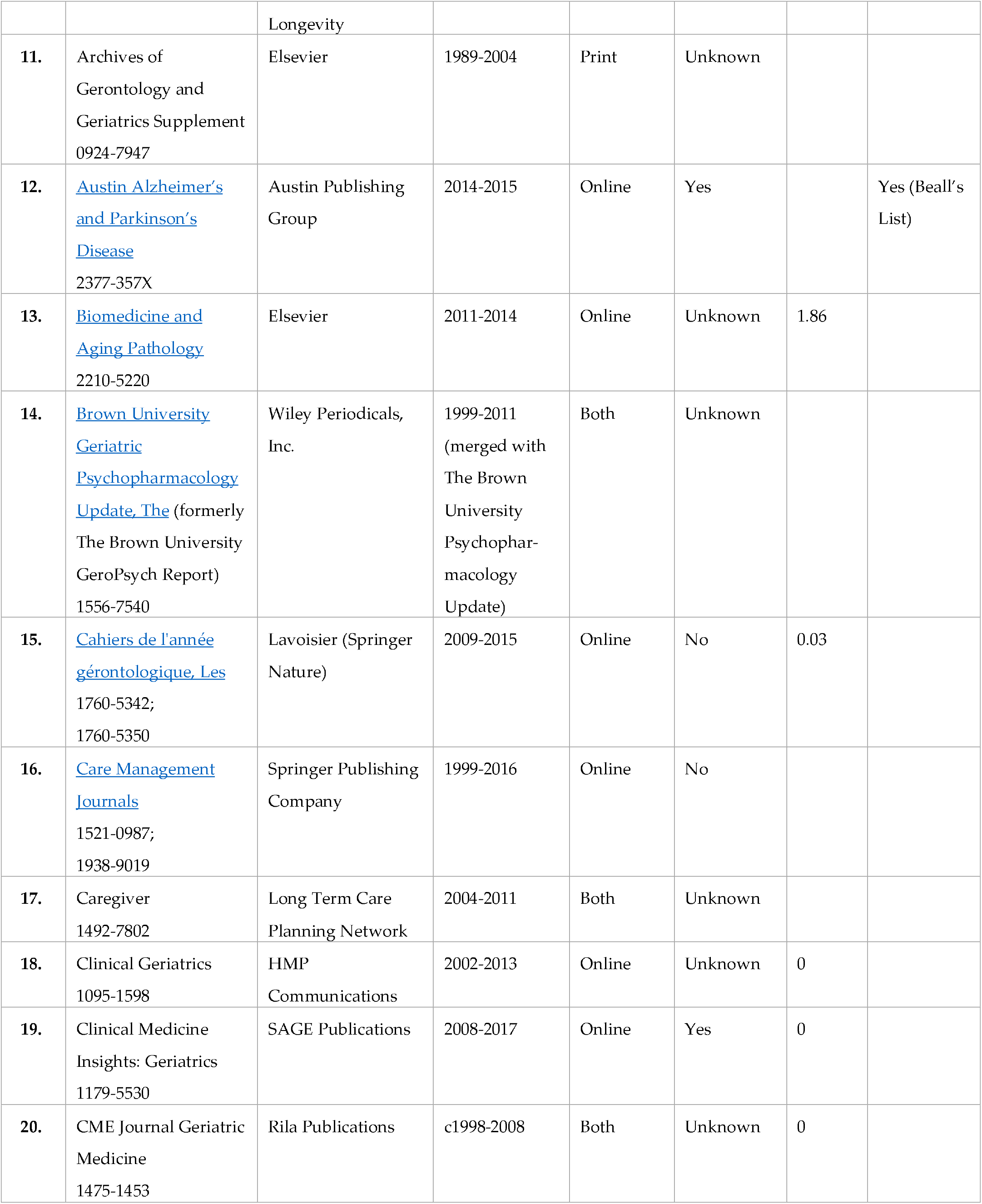
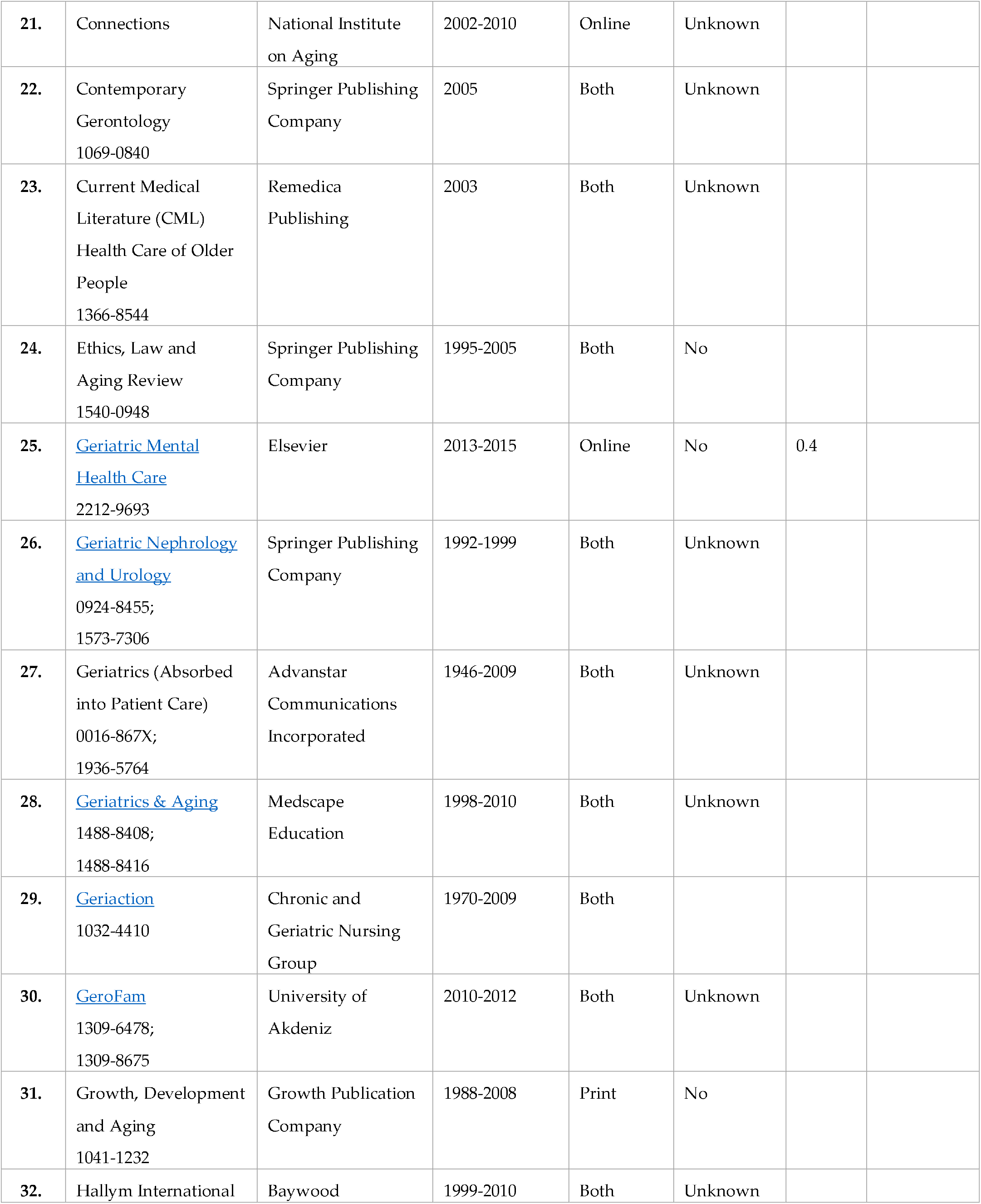
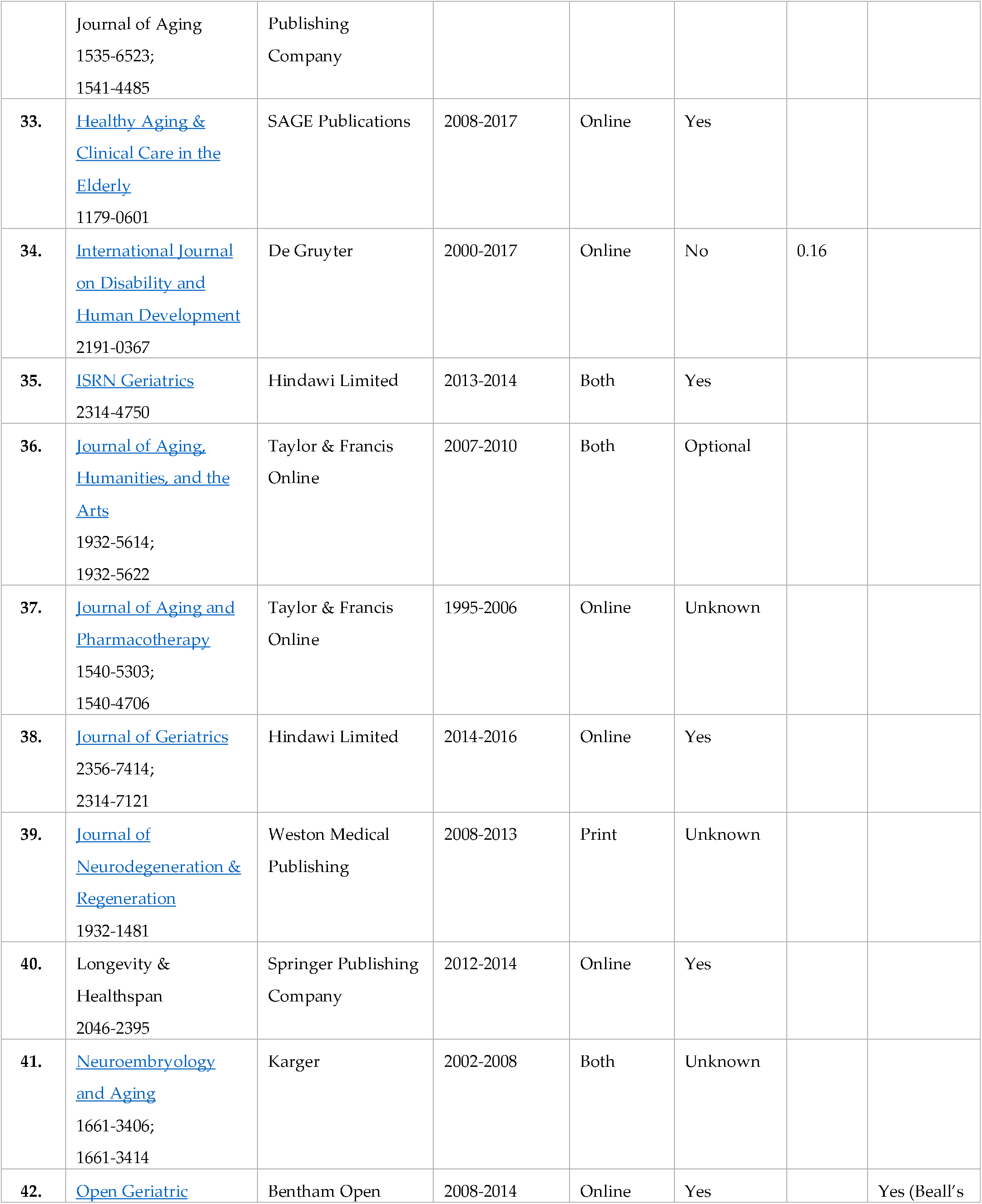
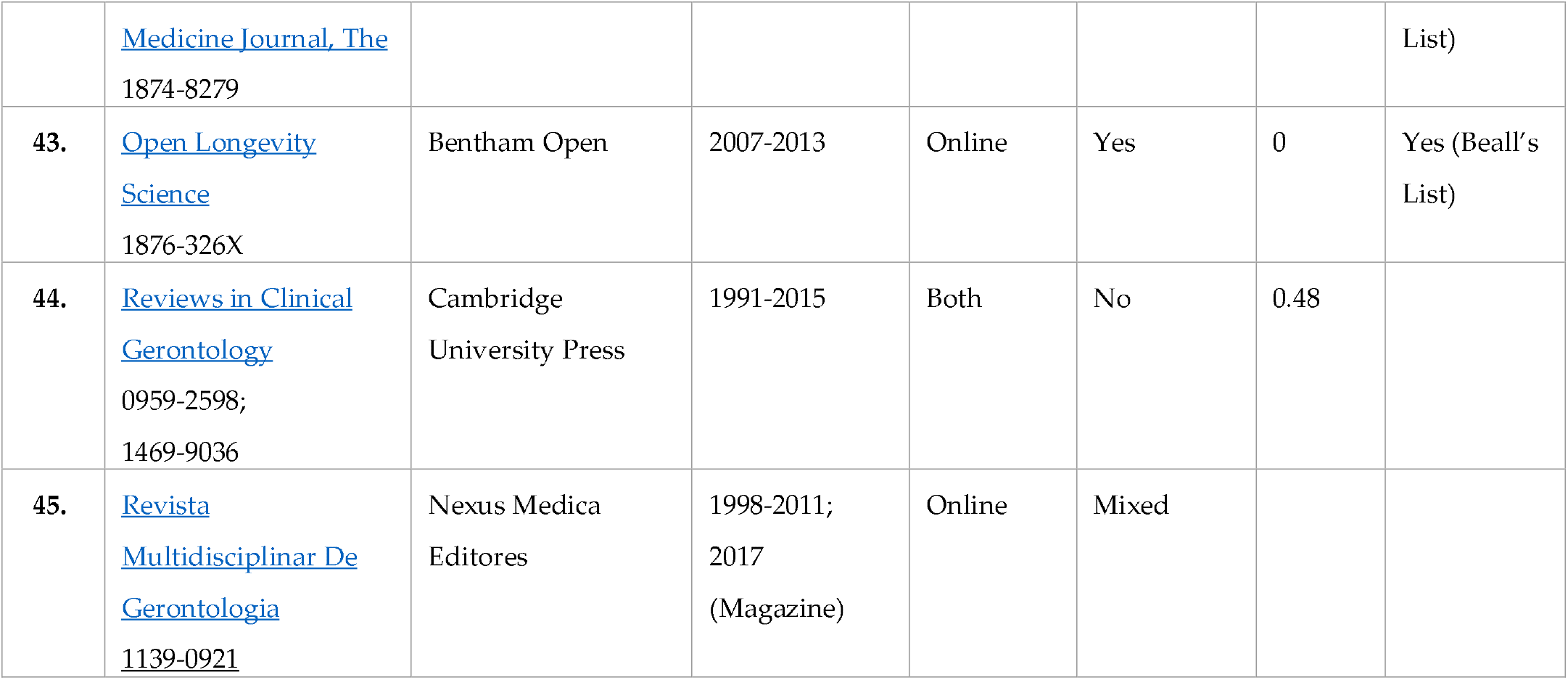
JOURNALS THAT HAVE CEASED PUBLICATION

| # | Name of Journal | Publisher/Society | Years of Publication | Print or Online? | Open Access? | Impact Factor | Predatory Publisher? |
| --- | --- | --- | --- | --- | --- | --- | --- |
| 1. | Advances in Cell Aging and Gerontology<br>1566-3124;<br>1875-5224 | Elsevier | 1996-2005 | Both | No |  |  |
| 2. | Advances in Geriatrics<br>2314-7679 | Hindawi Limited | 2014-2015 | Online | Yes |  |  |
| 3. | Ageing Horizons<br>1746-1073;<br>1746-1081 | The Oxford Institute of Population Ageing | 2004-2010 | Both | Yes |  |  |
| 4. | Ageing Research<br>2036-7384;<br>2036-7376 | PAGEPress | 2009-2012 | Online | Yes |  |  |
| 5. | Aging Health<br>1745-509X;<br>1745-5103 | Future medicine Ltd. | 2005-2013 | Both | Yes | 0 |  |
| 6. | Alzheimer's Care Today<br>1936-3001 | Wolters Kluwer | 2000-2010 | Both | Most likely optional |  |  |
| 7. | Alzheimer's Disease Research Journal<br>1935-2514 | Nova Science Publishers | Unknown | Print | No |  | Potentially |
| 8. | American Journal of Geriatric Cardiology, The<br>1751-715X | Wiley | 1992-2008 | Both | Most likely optional |  |  |
| 9. | American Journal of Geriatric Pharmacotherapy, The<br>1543-5946 | Elsevier | 2003-2013 | Print | Unknown |  |  |
| 10. | Approaches to Aging Control<br>1885-4028 | Spanish Society of Anti-Aging Medicine and | 2008-2015 | Online | Yes |  |  |
|  |  | Longevity |  |  |  |  |  |
| 11. | Archives of Gerontology and Geriatrics Supplement<br>0924-7947 | Elsevier | 1989-2004 | Print | Unknown |  |  |
| 12. | Austin Alzheimer's and Parkinson's Disease<br>2377-357X | Austin Publishing Group | 2014-2015 | Online | Yes |  | Yes (Beall's List) |
| 13. | Biomedicine and Aging Pathology<br>2210-5220 | Elsevier | 2011-2014 | Online | Unknown | 1.86 |  |
| 14. | Brown University Geriatric Psychopharmacology Update, The (formerly The Brown University GeroPsych Report)<br>1556-7540 | Wiley Periodicals, Inc. | 1999-2011<br>(merged with The Brown University Psychopharmacology Update) | Both | Unknown |  |  |
| 15. | Cahiers de l'année gérontologique, Les<br>1760-5342;<br>1760-5350 | Lavoisier (Springer Nature) | 2009-2015 | Online | No | 0.03 |  |
| 16. | Care Management Journals<br>1521-0987;<br>1938-9019 | Springer Publishing Company | 1999-2016 | Online | No |  |  |
| 17. | Caregiver<br>1492-7802 | Long Term Care Planning Network | 2004-2011 | Both | Unknown |  |  |
| 18. | Clinical Geriatrics<br>1095-1598 | HMP Communications | 2002-2013 | Online | Unknown | 0 |  |
| 19. | Clinical Medicine Insights: Geriatrics<br>1179-5530 | SAGE Publications | 2008-2017 | Online | Yes | 0 |  |
| 20. | CME Journal Geriatric Medicine<br>1475-1453 | Rila Publications | c1998-2008 | Both | Unknown | 0 |  |
| 21. | Connections | National Institute on Aging | 2002-2010 | Online | Unknown |  |  |
| 22. | Contemporary Gerontology<br>1069-0840 | Springer Publishing Company | 2005 | Both | Unknown |  |  |
| 23. | Current Medical Literature (CML)<br>Health Care of Older People<br>1366-8544 | Remedica Publishing | 2003 | Both | Unknown |  |  |
| 24. | Ethics, Law and Aging Review<br>1540-0948 | Springer Publishing Company | 1995-2005 | Both | No |  |  |
| 25. | Geriatric Mental Health Care<br>2212-9693 | Elsevier | 2013-2015 | Online | No | 0.4 |  |
| 26. | Geriatric Nephrology and Urology<br>0924-8455;<br>1573-7306 | Springer Publishing Company | 1992-1999 | Both | Unknown |  |  |
| 27. | Geriatrics (Absorbed into Patient Care)<br>0016-867X;<br>1936-5764 | Advanstar Communications Incorporated | 1946-2009 | Both | Unknown |  |  |
| 28. | Geriatrics & Aging<br>1488-8408;<br>1488-8416 | Medscape Education | 1998-2010 | Both | Unknown |  |  |
| 29. | Geriaction<br>1032-4410 | Chronic and Geriatric Nursing Group | 1970-2009 | Both |  |  |  |
| 30. | GeroFam<br>1309-6478;<br>1309-8675 | University of Akdeniz | 2010-2012 | Both | Unknown |  |  |
| 31. | Growth, Development and Aging<br>1041-1232 | Growth Publication Company | 1988-2008 | Print | No |  |  |
| 32. | Hallym International | Baywood | 1999-2010 | Both | Unknown |  |  |
|  | Journal of Aging<br>1535-6523;<br>1541-4485 | Publishing<br>Company |  |  |  |  |  |
| 33. | Healthy Aging & Clinical Care in the Elderly<br>1179-0601 | SAGE Publications | 2008-2017 | Online | Yes |  |  |
| 34. | International Journal on Disability and Human Development<br>2191-0367 | De Gruyter | 2000-2017 | Online | No | 0.16 |  |
| 35. | ISRN Geriatrics<br>2314-4750 | Hindawi Limited | 2013-2014 | Both | Yes |  |  |
| 36. | Journal of Aging, Humanities, and the Arts<br>1932-5614;<br>1932-5622 | Taylor & Francis<br>Online | 2007-2010 | Both | Optional |  |  |
| 37. | Journal of Aging and Pharmacotherapy<br>1540-5303;<br>1540-4706 | Taylor & Francis<br>Online | 1995-2006 | Online | Unknown |  |  |
| 38. | Journal of Geriatrics<br>2356-7414;<br>2314-7121 | Hindawi Limited | 2014-2016 | Online | Yes |  |  |
| 39. | Journal of Neurodegeneration & Regeneration<br>1932-1481 | Weston Medical<br>Publishing | 2008-2013 | Print | Unknown |  |  |
| 40. | Longevity & Healthspan<br>2046-2395 | Springer Publishing<br>Company | 2012-2014 | Online | Yes |  |  |
| 41. | Neuroembryology and Aging<br>1661-3406;<br>1661-3414 | Karger | 2002-2008 | Both | Unknown |  |  |
| 42. | Open Geriatric | Bentham Open | 2008-2014 | Online | Yes |  | Yes (Beall's) |
|  | Medicine Journal, The<br>1874-8279 |  |  |  |  |  | List) |
| 43. | Open Longevity Science<br>1876-326X | Bentham Open | 2007-2013 | Online | Yes | 0 | Yes (Beall's List) |
| 44. | Reviews in Clinical Gerontology<br>0959-2598;<br>1469-9036 | Cambridge University Press | 1991-2015 | Both | No | 0.48 |  |
| 45. | Revista Multidisciplinar De Gerontologia<br>1139-0921 | Nexus Medica Editores | 1998-2011;<br>2017<br>(Magazine) | Online | Mixed |  |  |

## 4. Discussion

The list provides clear indication that the print only journal is on its way out, with 7 currently published journals in print only, compared to 115 journals in both print and online formats, and 76 online only journals. We expect to see many of the journals still in print or both formats to move to online only within the next few years.

We are requesting that journal editors and publishers send updated information to the senior author to help improve the list and information about the journals. Missing journals need to be added to the list after they are identified. It is our expectation that updated versions of this paper may be published in a preprint form in the future. This can more easily be updated on a regular basis, or made available in some other format.

Of interest is the apparently low number of journals focusing on nursing in the field of gerontology. Only 7 of the 195 (4%) currently published journals specifically refer to nursing, a low number as it seems that most nurses do work with aging patients. Those in fields of sociology and social work as well as psychology seem under-represented. Another area of concern is the lack of journals publishing research focusing on caregiving, with the only journal we found in this area having ceased publication. The area of caregiving currently has an emphasis on problems of the health of caregivers and the problems associated with poor (even abusive) quality of care for the aged. This is also a field often staffed by a nonacademic populace who would not have easy access to the wealth of information potentially available on current gerontology and caregiving research.

It is apparent from a librarian’s point of view that it is difficult for the average researcher to find and compile a complete list of potential publishing outlets, and it would be helpful to the scholarly community to have an easily searchable database, preferably not behind a firewall. This could increase the diversity of journal articles being published and help researchers find more appropriate choices for article submission, as well as increasing the diversity of authors who are submitting works for publication.

With the rise in predatory publishing, we consider that having only 10% of the journals a low number, though there should be careful evaluation of all journals including newly launched titles. Additional journal titles are expected to be found as many of the predatory journals/publishers are not found in searches. More research could be done in specific scholarly fields as to the perceptions of predatory journals as well as how the rise of the open access movement has affected fields of research for better or for worse.

## 5. Conclusions

This paper provides a current update on journals being published in the broad disciplines of aging, geriatrics and gerontology and providing a broad view of research being published in the area. It is expected that this will be a useful resource to researchers who are considering publishing their research, to those trying to make sure they find all relevant articles in their searches, and for those looking at the discipline of aging, geriatrics and gerontology as a whole.

The data provide a view of the wide areas covered within the broad disciplines involved in aging, and the increasing numbers of journals does indicate both an increased focus on aging as well as the dynamic nature of the expanding aspects of aging. While many general journals publish articles on areas of aging, these specific journals are important as a way to view the activity and interests in these fields. While the increase in journal numbers and types is not a direct concern, it is critical that the field note the quality of journals, and that efforts are made to help other researchers avoid predatory journals where there are high costs but the quality of the reviews and papers are very low. This not only hurts the field and is a waste of money, but also undermines the research and medical communities and confidence in them.

This compiled list of data will hopefully be a jumping off point for other fields of research to compile lists and bibliographies of available journals for publication and their potential value or risk. It will be especially helpful to students and new researchers in the field as a starting point for their projects and careers.

## Data Availability

All data is available from author.

## Author Contributions

Conceptualization, R.W.; methodology, R.W.; investigation, H.B.; data curation, H.B.; writing—original draft preparation, H.B. and R.W.; writing—review and editing, H.B. and R.W.; visualization, H.B.; supervision, R.W.; project administration, R.W.

## Acknowledgments

Thanks for support and editing to Donna Wolff.

